# High vaccine coverage and non-pharmaceutical interventions decreased hospitalizations and deaths due to the Omicron variant in Newfoundland and Labrador, Canada

**DOI:** 10.64898/2026.11.02.26367081

**Authors:** Francis Anokye, Michael WZ Li, Amy Hurford

## Abstract

The Canadian province of Newfoundland and Labrador (NL) had high vaccination coverage, and few infections were reported prior to the establishment of the Omicron variant. We investigated whether changes in the timing and rate of booster dose administration could have reduced severe disease outcome and reliance on non-pharmaceutical interventions (NPIs) during the Omicron wave in NL. We used a vaccination-stratified epidemiological model calibrated to serology data to investigate the impact of vaccination relative to counterfactual vaccination strategies under actual, strict and no NPI scenarios. Compared with a Canadian vaccination rate scenario, the actual vaccination rollout in NL was estimated to have prevented approximately 519 hospitalizations and 122 deaths from December 15, 2021 to December 15, 2022, corresponding to reductions of 25.9% and 30.9%, respectively. These reductions reflected contributions from NL’s higher initial primary series coverage (81.3%), higher final booster dose coverage (57.3%), and the implementation of various NPIs for 89 days after the Omicron BA.1 variant became established. Earlier and faster administration of booster doses reduced the duration of strict NPIs required to achieve hospitalization targets. Our findings show how the timing and rate of vaccination rollout can complement NPIs to reduce severe disease and the duration for which strict NPIs are required.

## 1 Introduction

Early in the COVID-19 pandemic, non-pharmaceutical interventions (NPIs) were implemented to control the spread of the SARS-CoV-2 virus and consisted of social distancing, gathering limits, and business and school closures (Brauner et al, 2021; Flaxman et al, 2020; Sonabend et al, 2021). By mid-2021, a SARS-CoV-2 vaccine had been developed and distributed, and the global adult population was rapidly becoming vaccinated. Globally, COVID-19 vaccination was estimated to have prevented about 19.8 million deaths during the first year of vaccine deployment (Watson et al, 2022). In Canada, the implementation of NPIs and vaccination programs during the first two years of the pandemic reduced the number of pandemic deaths (Ogden et al, 2022). Despite their effectiveness, NPIs were economically and socially disruptive (Clemente-Suárez et al, 2021; Koren and Petö, 2020; Mofijur et al, 2021) and were ‘rolled back’ when local populations became highly vaccinated (Giordano et al, 2021; Moore et al, 2021). By mid to late 2021, many jurisdictions were in the process of ‘re-opening’, which involved implementing fewer and less restrictive NPIs (Brankston et al, 2022; Hale et al, 2021; Levitt et al, 2022), and the public was fatigued from prolonged periods of intermittent NPIs (World Health Organization, 2020).

After nearly two years of the pandemic, on November 24, 2021, the Omicron variant (B.1.1.529) was detected (Karim and Karim, 2021; Viana et al, 2022; World Health Organization, 2021), began spreading throughout the world, and was determined to have substantial immune evasion capabilities against both vaccine-derived and natural immunity (Shrestha et al, 2022). The Omicron variant was highly transmissible and likely to cause many severe infections (Nealon and Cowling, 2022; Shrestha et al, 2022; World Health Organization, 2021). An additional dose of vaccine, referred to as a ‘booster dose’, could decrease the chance of infection and the risk of severe disease due to infection with the newly emerged Omicron SARS-CoV-2 variant (Abu-Raddad et al, 2022; Andrews et al, 2022).

Prior to the establishment of the Omicron variant, cumulative SARS-CoV-2 incidence and vaccination coverage varied across provinces and territories in Canada (Maquiling et al, 2023; Murphy et al, 2023). The situation in Newfoundland and Labrador (NL) was distinctive due to the province’s low cumulative incidence and high vaccination coverage. Serosurveillance estimates show that around 2% of the NL population, as compared to 9% of the Canadian population overall, had experienced a SARS-CoV-2 infection by November 2021 (Murphy et al, 2023). This low cumulative incidence was due to several factors, including NL’s implementation of a containment strategy, such as travel measures (Mohammadi et al, 2025), and its enactment of a precautionary approach that protected vulnerable populations in the province (Government of Newfoundland and Labrador, 2022). By January 2022, NL was one of the most highly vaccinated jurisdictions in the world, with 93.7% of the NL population having received at least one dose of vaccine (Treble, 2022). This vaccination coverage included 74.7% of 5-11-year-olds, despite vaccinations becoming available to this age group only in mid-November 2021.

Whether alternative vaccination rollout strategies could have reduced the need for NPIs while mitigating the impact of the Omicron variant in NL is not known. We investigate the effect of different vaccination rollout and NPI implementation strategies specifically for Omicron variant spread in the Canadian province of NL. We focus on NL because the epidemiology and public health response to COVID-19 are regionally specific, vaccination and infection data are available for the NL population, and because we have estimated epidemiological quantities and reported key information for the COVID-19 pandemic in NL in our previous publications (Anokye et al, 2026; Doig et al, 2026; Hurford et al, 2023; Martignoni et al, 2023). Studying this population helps generalize recommendations for future pandemic preparedness.

We develop an epidemiological model to implement counterfactual scenarios where booster dose administration: began earlier; occurred at a slower or faster rate; or occurred as it did in the Canadian population. The Canadian rate scenario also assumes that the initial vaccination coverage is at either the NL or average Canadian levels. The effects of no NPIs, strict NPIs and the actual NPIs are assessed in combination with the different vaccination scenarios. We evaluate how vaccination affected the need for NPIs at the start of the Omicron wave in NL and determine the number of days that strict NPIs would need to be in place to achieve particular mitigation goals. We calculate the number of hospitalizations and deaths that would have occurred in NL under each of these counterfactual scenarios.

## 2 Methods

### 2.1 Model

We numerically solve a mechanistic Susceptible-Exposed-Asymptomatic-Infected-Recovered (SEAIR) model, previously developed by Anokye et al (2026), to quantify the impact of different combinations of vaccine administration and NPI implementation. Within this SEAIR structure (see Figure 1), susceptible individuals (*S*) become exposed (*E*) after contracting SARS-CoV-2 through contact with an infectious individual. The exposed individuals transition to either the asymptomatic (*A*) or symptomatic (*I*) compartments after a latent period. These individuals later recover and move on into the recovered compartment (*R*) where they remain immune to reinfection.

**Fig. 1.**
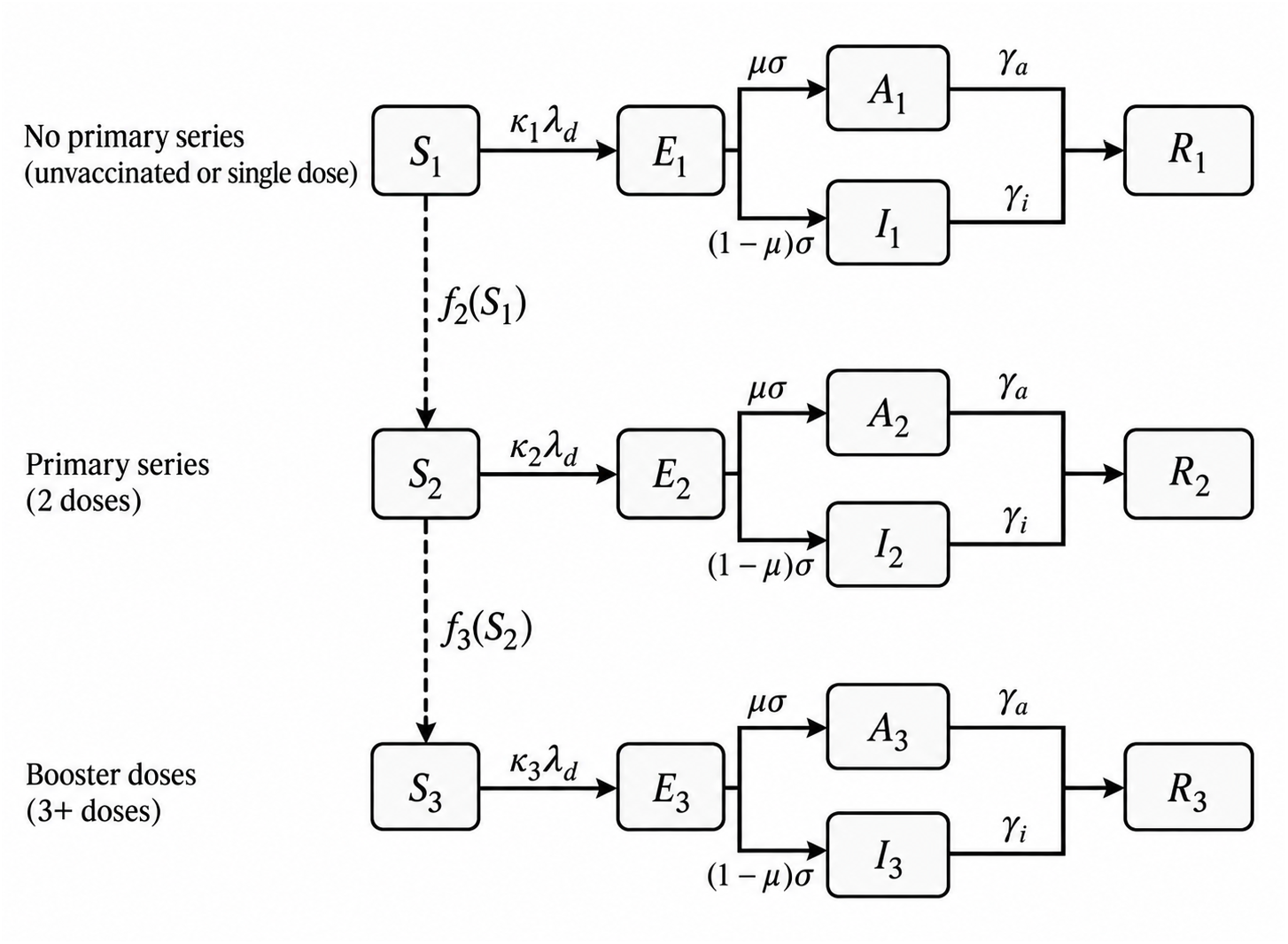
Vaccination-stratified SEAIR model. The population is divided into three vaccination cohorts: no primary series, primary series, and booster doses. Each cohort includes susceptible, exposed, asymptomatic infectious, symptomatic infectious, and recovered compartments. Solid arrows indicate transitions associated with infection and recovery, while dashed arrows represent movement between susceptible compartments due to vaccination.

Our model stratifies the population of NL into three cohorts based on vaccination status after receiving a Health Canada-approved COVID-19 vaccine, without distinguishing between vaccine types. The cohorts include unvaccinated individuals or those who have received one dose from a two-dose primary series (referred to as ‘no primary series’), individuals who have completed the primary series (referred to as ‘primary series’), and those who have one or more additional doses after receiving the primary series (referred to as ‘booster doses’). Collectively, the ‘no primary series’ and ‘primary series’ cohorts are referred to as ‘no booster doses’. Individuals can transition from the no primary series cohort to the primary series cohort, and then from the primary series cohort to the booster dose cohort. This assumption matches the definition of these vaccination statuses, for example that individuals must complete the primary vaccination series before receiving a booster dose.

The protection conferred by vaccination is modelled as a reduction in susceptibility to infection, and this is occurs through cohort-specific parameters (*κ*_1_, *κ*_2_, and *κ*_3_) that contribute to the force of infection. The model assumes vaccine-induced protection is immediate. The original model diagram and parameterization are provided in Anokye et al (2026). We assume only susceptible individuals in each vaccination cohort can receive vaccine doses and transition to another vaccination cohort. We do not model vaccination for exposed, asymptomatic, or symptomatic individuals since few individuals with these states are likely to get vaccinated. We do not model vaccination of recovered individuals because recovered individuals are assumed to be immune from infection, such that vaccination does not afford additional protection, and modelling the vaccination of recovered individuals is not likely to change our results.

Hospital admissions and deaths were calculated directly from infection counts using vaccination-specific perinfection severity risks. We let *I*_1,*t*_, *I*_2,*t*_ and *I*_3,*t*_ denote the number of new infections on day *t* among individuals having received no primary series, a primary series, and a booster dose. The daily number of hospital admissions, *H*_*t*_, and deaths, *D*_*t*_, were calculated as:

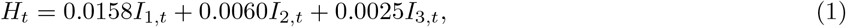

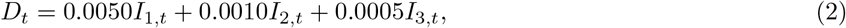

where the probabilities of hospitalization given infection were 0.0158, 0.0060 and 0.0025 for individuals with no primary series, a primary series, and a booster dose, respectively. Given the lack of an infection-hospitalization risk estimates specific to NL, these probabilities were estimated assuming that 0.58% of infections resulted in hospitalization, which corresponds to the lower bound of an estimate reported during the Omicron period in England (0.62%, 95% CI: 0.58–0.65%; Ward et al 2024), and due to the relative risks for each vaccination group (Danza, 2022; Havers et al, 2022). The probability of death given infection is 0.005 (Danza, 2022), 0.001 (Buchan et al, 2022; Danza, 2022) and 0.0007 (Buchan et al, 2022; Danza, 2022; Grewal et al, 2023) for the respective cohorts.

The initial conditions for our simulations are the model variable values on December 15, 2021 and were determined by implementing a burn-in period of 150 days to enable the number of individuals in each state to approach equilibrium proportions. The results we report consider the value of model variables from December 15, 2021 to December 15, 2022. Our model is parameterized for the BA.1 Omicron variant. During the COVID-19 pandemic, the BA.2 variant replaced the BA.1 variant after a few months in NL. Our simulations aim to understand the long-term impact of vaccination and NPI implementation on BA.1 spread under the hypothetical situation whereby BA.1 was not replaced by any other variants. We assume no waning of immunity and the simulation period we have chosen is sufficiently long such that herd immunity is surpassed and any further infection that occurs contributes negligibly to the total number of infections. The full system of equations and parameter estimates for our model are provided in the Supplementary Material.

#### NPI scenarios

NPI implementation in NL during the spread of the Omicron variant consisted of an alert level system and delayed re-opening of kindergarten to grade 12 schools as described in (Anokye et al, 2026). The public health emergency was repealed in NL on March 14, 2022 (Government of Newfoundland and Labrador, 2022) and at this time NPI implementation to mitigate the spread of COVID-19 returned to pre-pandemic levels. To estimate the actual time-varying transmission rate of SARS-CoV-2 from December 15, 2021 to March 14, 2022, we refitted our model to serology data following the approach described in Anokye et al (2026). The transmission rate, *β*(*t*), was modelled as a smooth exponential spline using natural cubic spline basis functions, with the spline coefficients estimated by fitting the model to seroincidence data. Further details on the formulation and fitting procedure are provided in Anokye et al (2026). The model fit to the seroincidence data (Fig. S1) and estimated trajectory of *β*(*t*) (Fig. S2) are shown in the Supplementary Material.

For all NPI scenarios, for March 15, 2022–December 15, 2022, we set the transmission rate equal to the maximum value estimated during the period December 15, 2021–March 14, 2022 (*β*(*t*) = 0.3675; *R*_*t*_ = 2.43). This allows us to compare NPI scenarios. For December 15, 2021–March 14, 2022, the scenarios are: (1) Actual NPIs: the time-varying transmission rate as estimated from the NL serology data; (2) No NPIs: the maximum of time-varying transmission rate as estimated for the actual NPIs scenario (*β*(*t*) = 0.3630; *R*_*t*_ = 2.40); (3) Strict NPIs: the minimum of the time-varying transmission rate as estimated for the actual NPIs scenario (*β*(*t*) = 0.2467; *R*_*t*_ = 1.63). These scenarios are shown in Figure 2.

**Fig. 2.**
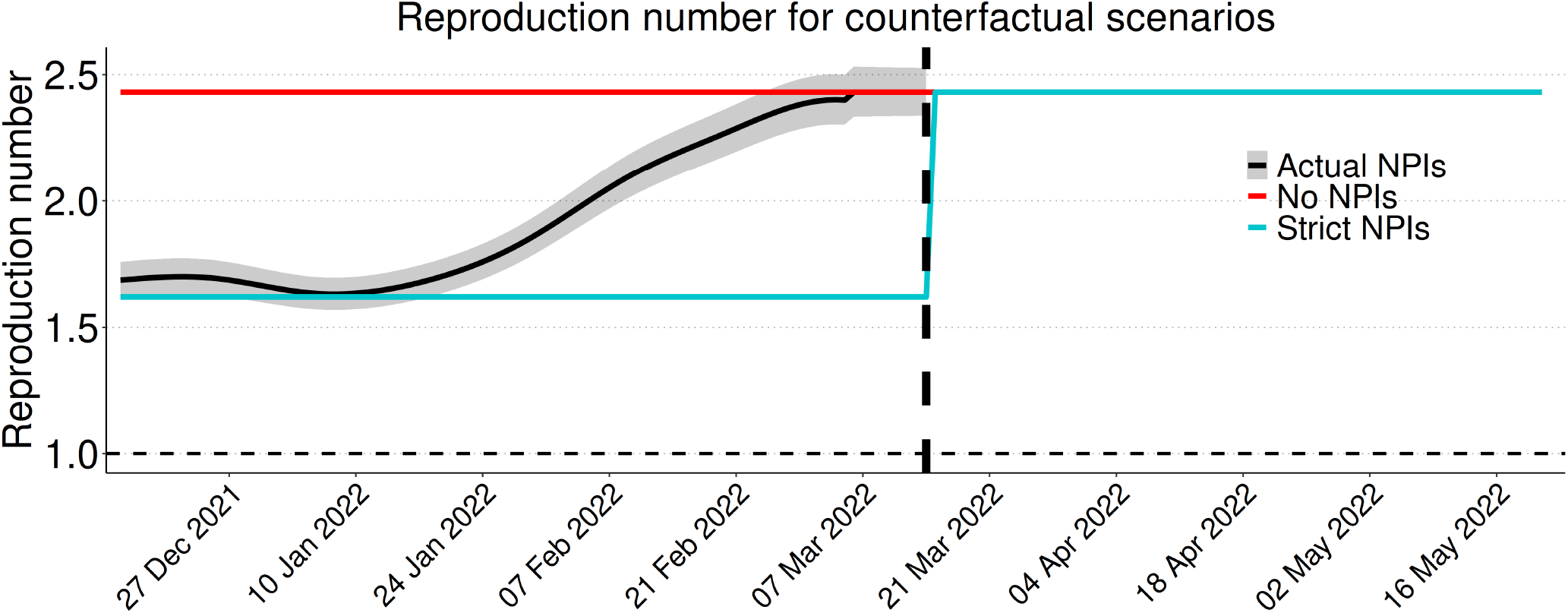
NPI scenarios for Omicron transmission in NL. The effective reproduction number over time for the actual NPI (solid black line with 95% CIs shown with the grey shaded region), no NPI (red line), and strict NPI (teal line) scenarios. The vertical dashed line indicates when NPIs were removed for all scenarios, and the horizontal dashed line shows the epidemic threshold of 1.

We also consider how long strict NPIs (defined as the minimum transmission rate estimated during the serology data fitting; *β*(*t*) = 0.2467; *R*_*t*_ = 1.63) would need to be implemented to achieve 1254 total hospitalizations and 28 peak daily hospital admissions for the period December 15, 2021–December 15, 2022. For these simulations, we consider the actual booster dose scenario and strict NPI implementation beginning on December 15, 2021 and lasting for a given number of days.

#### Vaccination scenarios

##### The ‘actual’ vaccination scenario

In NL, booster doses were first offered on November 8, 2021 to people for whom at least six months had passed since their second dose (Government of Newfoundland and Labrador, 2022). Most of the NL population was ineligible at this time as only 9,673 people (*≈* 1.9% of the NL population) met the necessary condition of at least 6 months since their second dose (Public Health Agency of Canada, 2026). The administration of booster doses in NL accelerated substantially 35 days later (beginning December 13, 2021) when the required interval between receiving two vaccine doses was decreased to 22 weeks. This change in eligibility meant that 154,100 people (*≈* 30% of the NL population) were now eligible for booster doses.

We obtained weekly cumulative vaccination counts for NL and Canada from the Public Health Agency of Canada website (Public Health Agency of Canada, 2022, 2026). These records include individuals who completed the primary (one or two dose) series and those who received additional booster doses. First and second additional doses administered after the primary series were combined into a single booster category.

The vaccination data for NL had missing records for booster doses from the start date, 8 November 2021 to 7 February, 2022. We interpolate these missing values by considering a report from the Newfoundland and Labrador Department of Health and Community Services (Government of Newfoundland and Labrador, 2022), which stated that approximately 25% of eligible individuals in the province had received a booster dose by January 11, 2022 (Government of Newfoundland and Labrador, 2022). Therefore, 20.9% of the total NL population had received a booster doses by January 11 (see Figure 3B; marked as a red star) because the eligible population was individuals aged 12 years and older (426,198 individuals) and the total population of NL is 510,550 individuals. We used this information and applied a monotonic power-law function to interpolate and fill the missing booster dose administration records (Figure 3B; black triangles). We required that the interpolated booster coverage rose gradually to reach 20.9% of the total population on January 11, 2022 and then continued to increase until February 7, 2022.

**Fig. 3.**
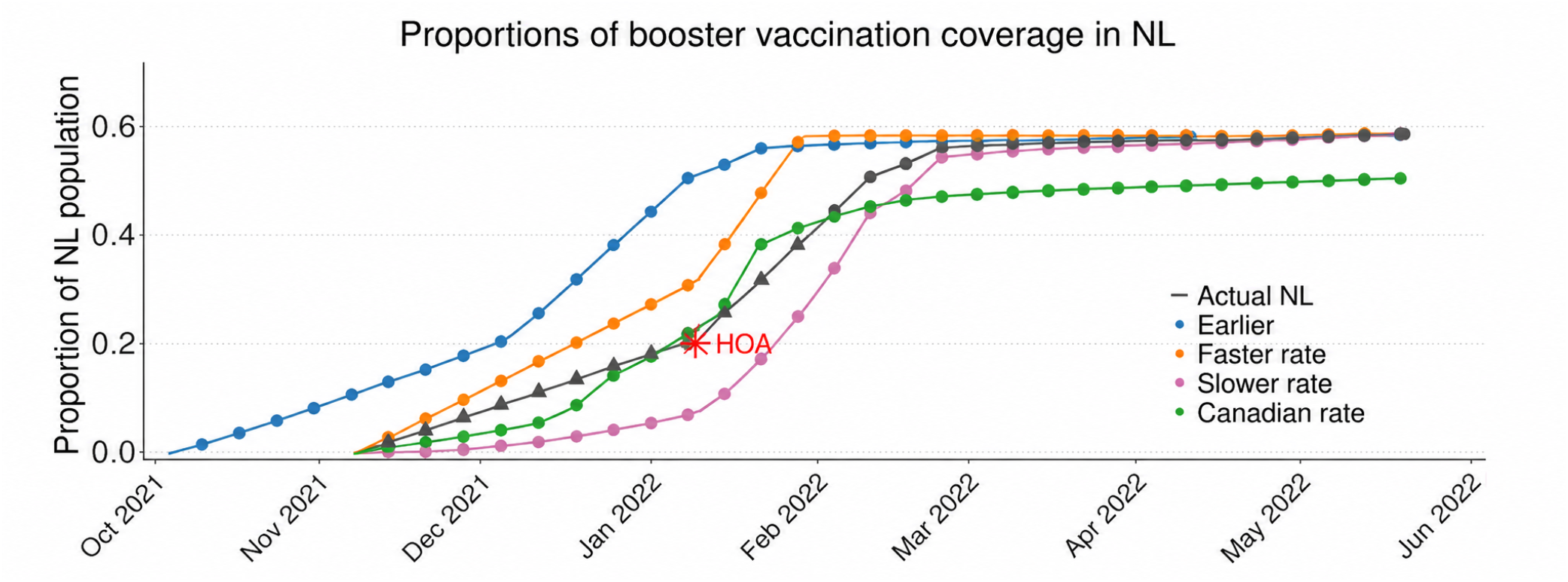
Booster vaccination scenarios for Omicron transmission in NL. The actual booster dose administration (black line) consists of values reported from PHAC data (dots; (Public Health Agency of Canada, 2024)) and interpolated estimates (triangles) to fill in missing data, where the interpolation was performed to be consistent with the estimated value of 20.9% of the population having received booster doses as of January 11, 2022 (see Section 2.1). The counterfactual scenarios are booster doses administered: earlier (blue), at a slower rate (magenta); at a faster rate (orange); and at the rate for the Canadian population.

Vaccination count data were reported as weekly aggregates and had constant values at some periods followed by sudden increases, which may be due to delayed reporting. We smoothed the counts by redistributing later increases backward across the constant periods to correct the gaps and delayed reporting. We converted these corrected weekly cumulative counts into a daily cumulative vaccination series using a monotone cubic-spline interpolation with an Hyman filter, and calculated daily rates as first differences of the interpolated cumulative series. The resulting interpolated data is used for the ‘actual’ scenario for vaccination in NL and is shown in Figure 3 (black line) alongside the PHAC vaccination data (black dots).

##### Counterfactual vaccination scenarios

The booster vaccination scenarios for NL were constructed by varying the timing and rate of booster administration, as well as a ‘Canadian rate’ scenario (Figure 3B; green line). Under the ‘slower rate’ booster scenario (Figure 3B; purple line), booster doses were administered at 50% of the actual NL booster dose administration rate. Under the ‘faster rate’ booster scenario (Figure 3B; orange line), booster doses are administered at 150% of the actual NL rate. In the ‘earlier’ booster scenario (Figure 3B; blue line), the actual booster administration curve is shifted to occur 35 days earlier. Thirty-five days was selected since this is the number of days between when booster dose administration first occurred in NL (but with restricted eligibility) and when eligibility requirements were relaxed (see Section 2.1). For all of the ‘slower rate’, ‘faster rate’, and ‘earlier’ booster dose scenarios, the asymptotic proportion of the population that receives booster doses is 57.3 % and is equal to that of the ‘actual’ scenario, which reflects the PHAC vaccination data. In addition, for all of these scenarios, the proportion of the population in each vaccination cohort when the booster dose rollout begins is equal to that of the ‘actual’ scenario, although we note that this start date is 35 days earlier for the ‘earlier’ scenario.

To construct the ‘Canadian rate’ scenario, we assumed that booster vaccination in NL followed the Canadian national average rather than the observed NL rollout that occurred. Using the PHAC vaccination data for November 6, 2021, we calculated the proportion of individuals in Canada who had one or fewer vaccine doses, had completed the primary vaccination series, and had received booster doses, and applied this proportion to the NL population to estimate the initial number of individuals in the no primary series, primary series, and booster dose vaccination cohorts. These calculations were used to determine the rates of transition between the cohorts and the Michaelis-Menten method (Bolker, 2008) was applied to preserve the non-negativity of model variables. For the Canadian rate scenario, booster dose coverage in NL asymptotes to 53.1 % of the total population and the initial number of individuals in the vaccine cohorts is set to the Canadian values. Figure 3B shows booster dose administration for all scenarios, and Table 1 notes other differences (i.e., initial vaccination coverage) and provides summary statistics.

**Table 1.** Additional information on the vaccination scenarios. The rate of booster dose administration for each of the vaccination scenarios is shown in Figure 3B. Additionally, these scenarios can differ in the start date of the booster dose administration program, the percentage of the population that at the start date have: not completed the primary series; have completed only the primary series; and have received one or more booster doses, and the final proportion of the total population that receives booster doses. Booster rollout rates are defined relative to the actual NL rollout, except for the Canadian rate scenario, which follows the Canadian national rollout.

| Comparison | Scenario | Booster start date | Initial vaccination coverage (%)<br>(No primary / Primary only / Booster) | Booster rollout rate | Final booster coverage (%) |
|---|---|---|---|---|---|
| Actual | Actual NL | Nov. 8, 2021 | 17.1 / 81.3 / 0.0 | Actual NL | 57.3 |
|  | Canadian rate | Nov. 8, 2021 | 22.3 / 76.7 / 0.2 | Canadian | 53.1 |
| Speed | Slower rate | Nov. 8, 2021 | 17.1 / 81.3 / 0.0 | 0.5× Actual NL | 57.3 |
|  | Faster rate | Nov. 8, 2021 | 17.1 / 81.3 / 0.0 | 1.5× Actual NL | 57.3 |
| Timing | Earlier | Oct. 4, 2021 | 22.9 / 75.6 / 0.0 | Actual NL (earlier start) | 57.3 |

We completed additional simulations to determine if differences between the Canadian rate and actual NL scenarios were due to: lower initial primary series coverage in Canada as compared to NL (see Table 1); or differences in booster dose administration in NL (black line) versus Canada (green line; Figure 3). We compared four scenarios: (i) Canadian initial vaccination coverage with Canadian vaccination rates (‘Canadian rate’); (ii) NL initial vaccination coverage with Canadian vaccination rates (‘Canadian rate (NL initial)’); (iii) Canadian initial vaccination coverage with the actual NL vaccination rates (‘Actual NL (Canadian initial)’); and (iv) NL initial vaccination coverage with the actual NL vaccination rates (‘Actual NL’). For scenarios sharing the same initial coverage, the numbers of individuals in the no primary series, primary series, and booster dose cohorts were identical at initialization and vaccination rates were implemented as per-capita transition rates applied to the number of eligible individuals in the corresponding vaccination compartment.

## 3 Results

Across all vaccination scenarios, strict NPI implementation led to delayed and reduced epidemic peaks and prolonged outbreaks (Figure 4 right column), while the absence of NPIs resulted in earlier and larger epidemic peaks with shorter outbreaks (Figure 4 left column). Across all NPI scenarios, a faster rate (Figure 4A-C) and earlier start (Figure 4D-F) of booster dose vaccination (as compared to the NL rate), and the actual NL vaccination rollout (as compared to the Canadian average rate; Figure 4G-I) delayed and reduced epidemic peaks and prolonged outbreaks. In Figure 4, the incidence in individuals who had not received booster doses is shown with light shading. The corresponding cumulative number of infections occurring in individuals without booster doses across these vaccination and NPI scenarios is shown in Fig. S3 in the Supplementary Material. When infections occur only after individuals have received booster doses, the probability of hospitalization (equation 1) and death (equation 2) decreases, which we explore in the following paragraphs.

**Fig. 4.**
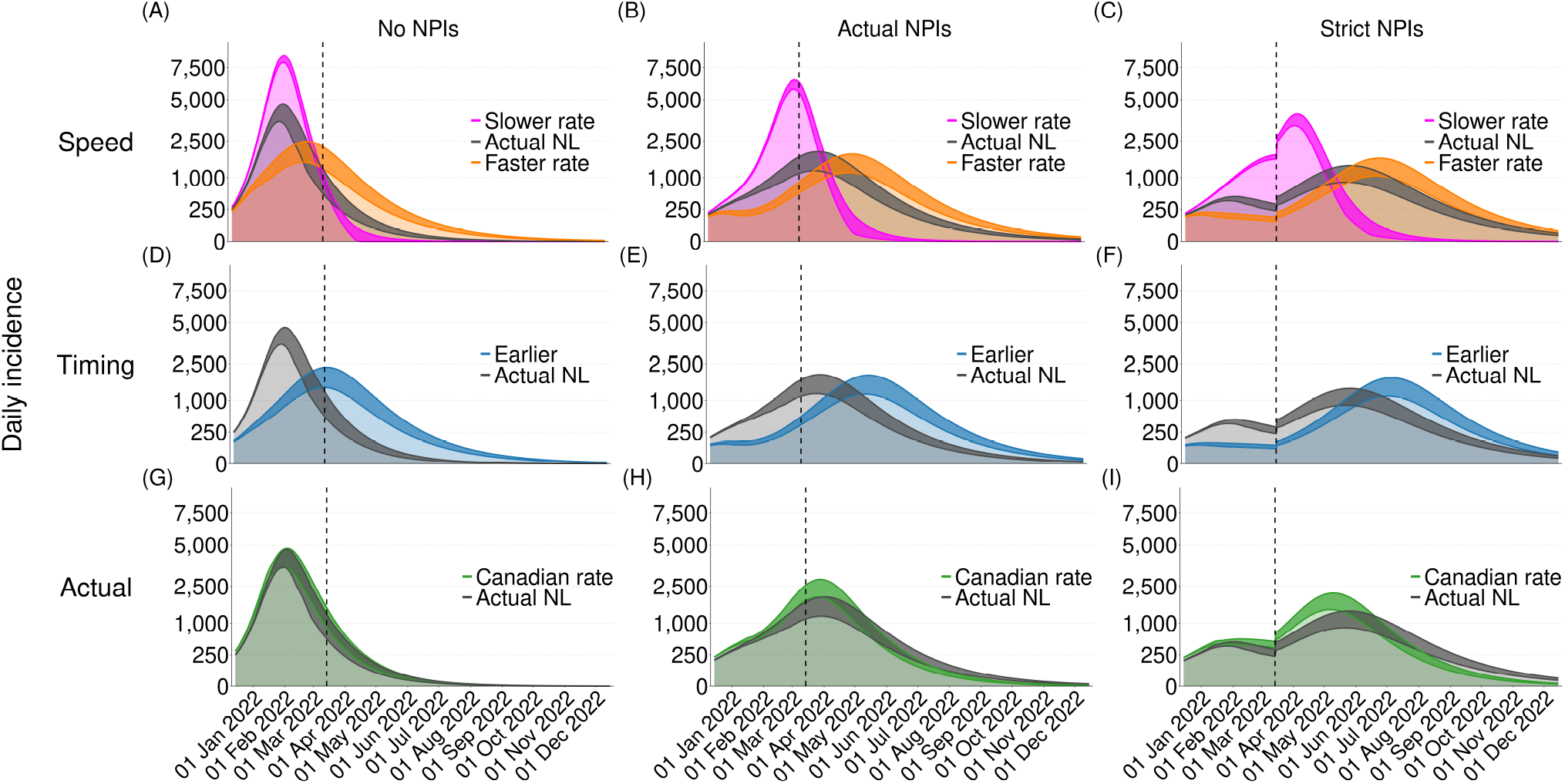
Strict NPIs and earlier and faster booster dose administration delay infections and reduce the epidemic peak. The temporal dynamics of infections among individuals with no booster dose (light shading) and booster doses (dark shading) are shown for no NPIs (left column), actual NPIs (middle column), and strict NPIs (right column). The effects of booster dose administration speed are compared for the slower, actual NL, and faster rates (A–C); timing is compared for the earlier and actual NL scenarios (D–F); and the Canadian rate is compared with the actual NL rate (G–I). The vertical dashed line indicates when all public health restrictions were removed in NL. The vertical axis has been square-root transformed to support visualization of the key effects.

When infections were delayed, fewer people were infected before receiving a booster dose (Fig. S3; Supplementary Material), which corresponded to fewer hospitalizations and deaths (Figure 5). For all vaccination scenarios, strict NPIs resulted in fewer hospitalizations and deaths than actual NPIs and no NPIs (Figure 5C compared with A and B). For the no NPI scenario, the ranking of the vaccination strategies (from highest hospitalizations and deaths to lowest) was slower rate, Canadian rate, then actual NL rate, with the earlier and faster rate scenarios performing similarly and best (Figure 5A). For the actual and strict NPI scenarios (Figure 5B), these relative rankings of the vaccination strategies remain the same, except that the actual NL scenario performs more similarly to the earlier and faster scenarios, which are still the best strategies (Figure 5B–C). Figure 5 suggests that the combination of the NPIs and booster dose vaccination rollout that actually occurred in NL during the early spread of the Omicron variant was effective in reducing hospitalizations and deaths, but this efficacy was contingent on the implementation of NPIs, as otherwise the NL vaccination rollout would have been outperformed by an earlier or faster vaccination rollout.

**Fig. 5.**
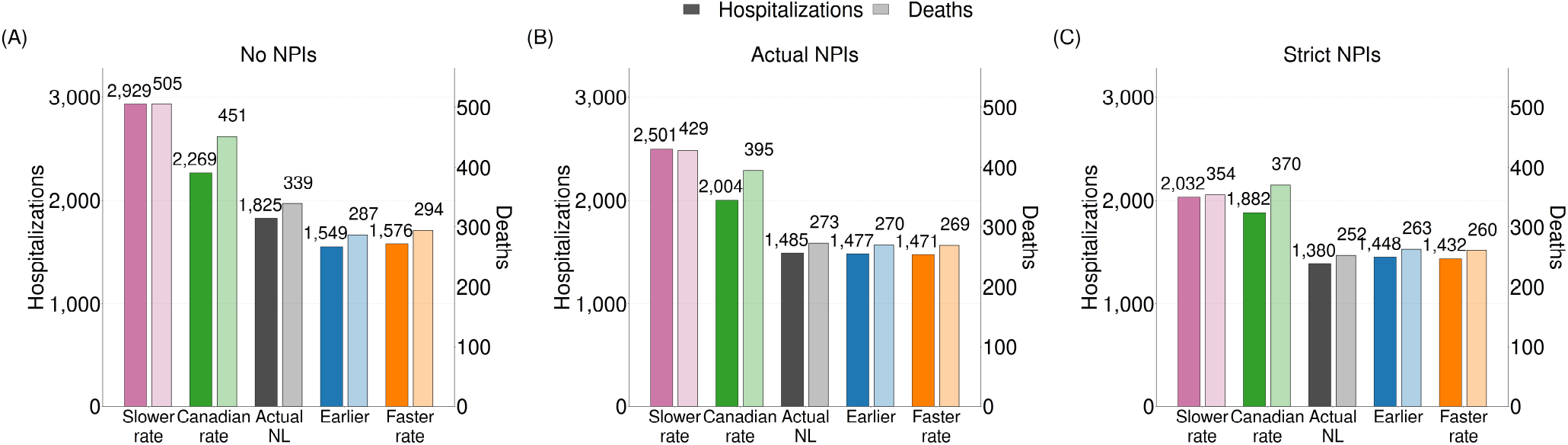
Strict NPIs and differences in the speed, timing, and actual vaccination rollouts reduce the total number of hospitalizations and deaths (December 15, 2021–December 15, 2022). When strict NPIs are implemented (December 15, 2021–March 14, 2022) the total number of hospitalizations and deaths is reduced compared with actual NPIs and no NPIs (C as compared to A and B). Faster and earlier administration of booster doses also reduces the total number of hospitalizations and deaths compared with the actual NL vaccination rollout. The actual NL vaccination rollout reduces the total number of hospitalizations and deaths compared with the Canadian vaccination rollout scaled to the NL population.

To understand the reduced number of hospitalizations and deaths (Figure 5D-F) for the actual NL (black bars) as compared to the Canadian average (green bars) scenarios, we consider scenarios with the initial NL vaccination coverage where booster doses were implemented at the Canadian rate (Figure 6A-C, yellow lines) and the initial Canadian vaccination coverage where booster doses were implemented at the NL rate (Figure 6A-C, dark green line). Both of these scenarios (yellow bars and dark green bars), which contained one element of the actual NL and Canadian average vaccination scenarios, performed intermediate to the actual NL (black bars) and Canadian average scenario (Figure 6D-F, which shows reductions relative to the Canadian average scenario, i.e., see Figure 5D-F green bars). Both the higher initial primary series coverage in NL prior to the administration of booster doses (yellow bars) and the higher final booster dose proportion (dark green bars) contributed to reducing the number of hospitalizations (by 10.1% and 15.1%, respectively) and deaths (by 17.2% and 12.9%, respectively) that occurred in NL (Figure 6E). The effect of NL’s higher initial primary series coverage and higher final booster dose coverage is nearly additive. Figures 5E and 6E show that our modelling predicts that NL’s high vaccination coverage reduced the number of hospitalizations by 519 (25.9%) and the number of deaths by 122 people (30.9%) that occurred in NL from December 15, 2021–December 15, 2022, relative to vaccination if it had occurred at the Canadian rate under the actual NPI scenario. Therefore, both NL’s higher initial vaccination coverage and higher percentage of people that received booster doses were important components of reducing hospitalizations and deaths.

**Fig. 6.**
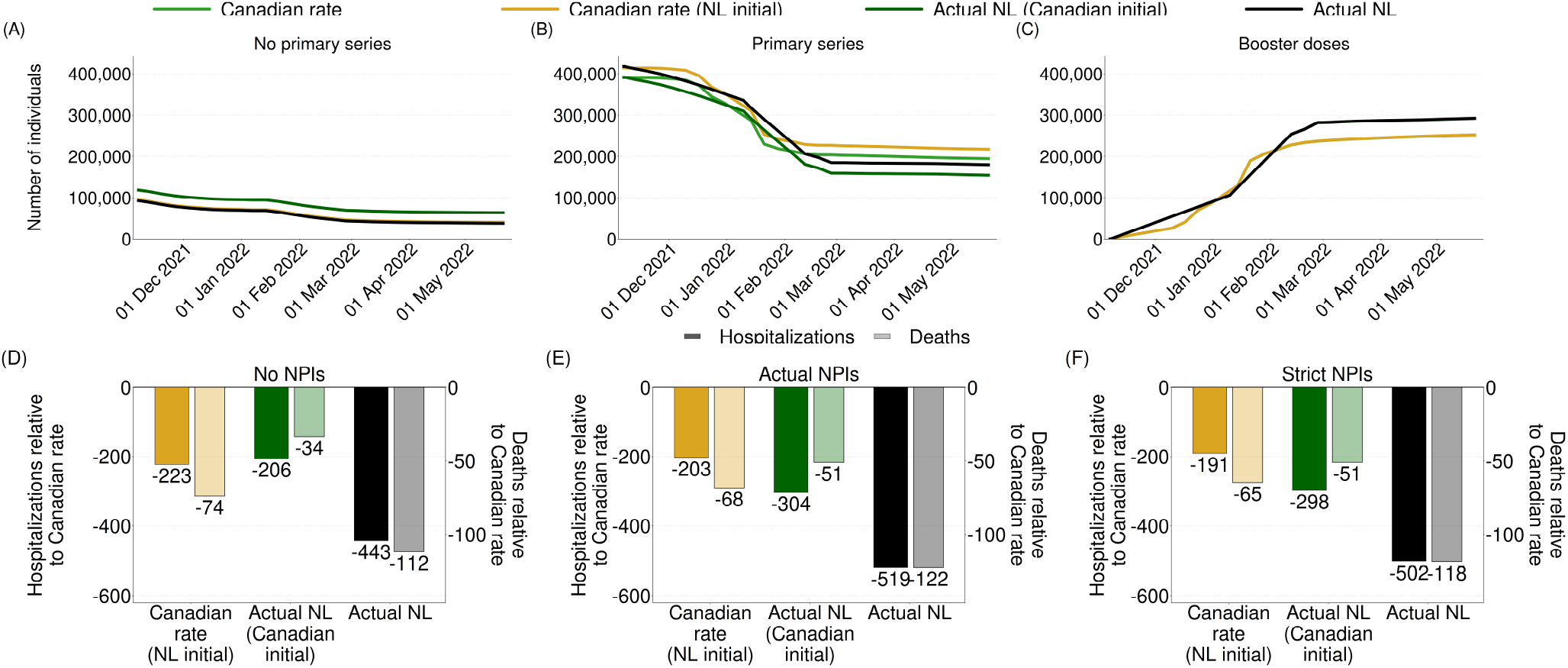
Both higher primary series coverage and higher booster dose coverage in NL relative to the Canadian average reduced hospitalizations and deaths (December 15, 2021–December 15, 2022). A-C The number of individuals with different vaccination statuses given different initial coverage and booster dose administration strategies (colors). D-F Number of hospitalizations (dark shading) and deaths (light shading) given no NPIs, actual NPIs, and strict NPIs for the different vaccination scenarios.

For our three best-performing booster dose administration strategies, 43–57 days of strict NPI implementation are sufficient to meet the target number of total (A) and peak (B) hospitalizations (Figure 7 blue, orange, and black lines). For a less stringent target, i.e., 1,700 total hospitalizations or a peak number of 25 daily hospital admissions, for the early (blue) and faster (orange) booster dose administration strategies, these targets could be met without any strict NPIs. In NL, some form of NPIs were implemented from December 15, 2021–March 14, 2022 (89 days), although these NPIs were often weaker than our definition of strict, and so we find that 56–57 days of strict NPIs would be sufficient to meet the hospitalization targets for the actual NL vaccination scenario (Figure 7A,B; black line). The number of days that strict NPIs needed to be implemented in NL to achieve these targets could be reduced by 7–14 days if booster doses had been implemented earlier or more quickly, where the exact number of days reduction depends on whether it is necessary to meet both hospitalization targets. Conversely, if vaccination coverage in NL was instead that of the Canadian average (Figure 7A,B; green line) the number of days that strict NPIs needed to be implemented to achieve the hospitalization targets increases from 56–57 days to 101–147 days. We found that even with 150 days of strict NPIs it was not possible to achieve the target number of peak hospitalizations if booster doses were administered at the slow rate (7B pink line).

**Fig. 7.**
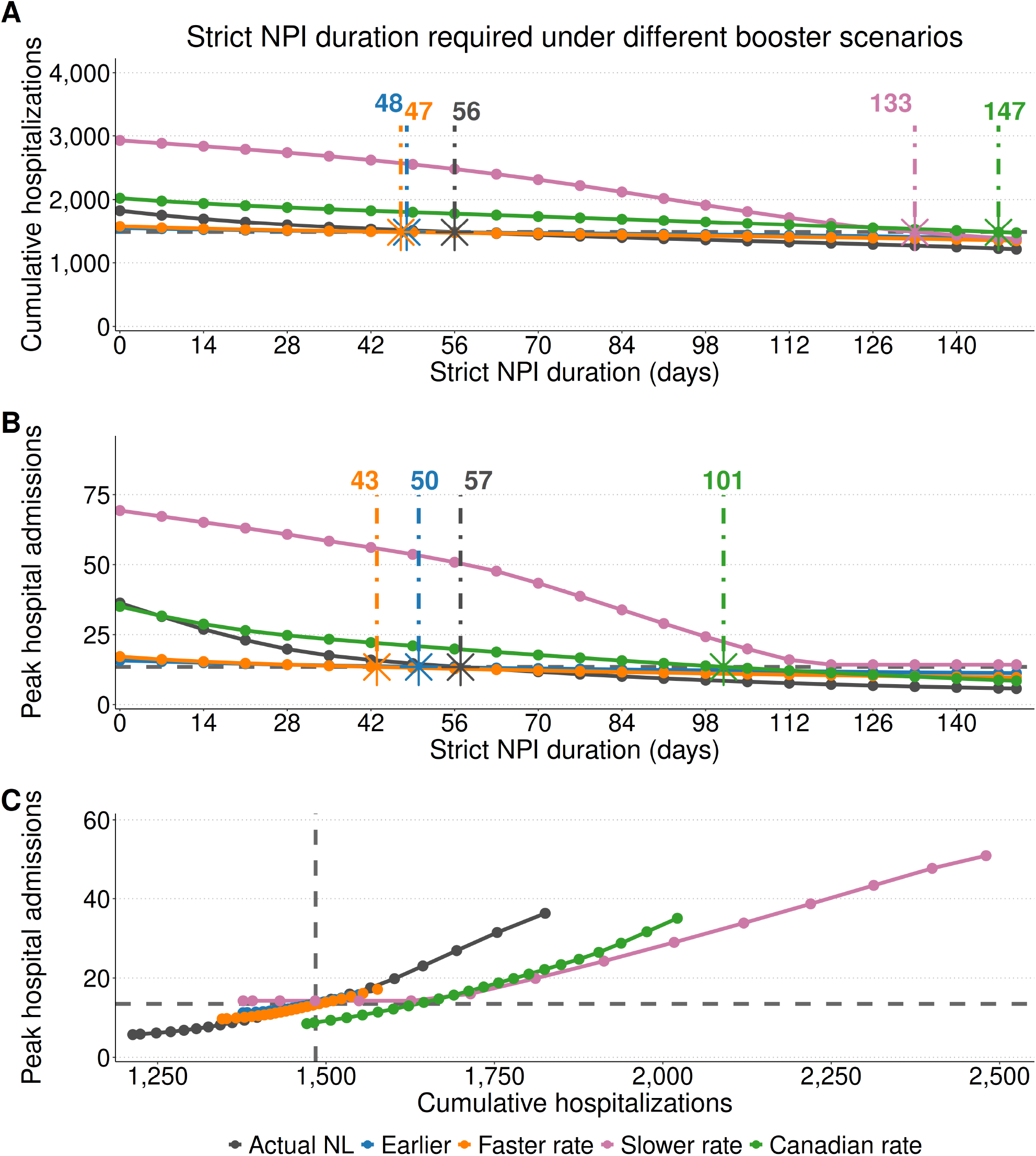
Earlier and faster booster dose administration reduced the duration of strict NPIs required to meet hospitalization targets. For the best-performing booster dose administration scenarios, 43–57 days of strict NPI implementation are sufficient to meet hospitalization targets. (A) Cumulative hospitalizations and (B) peak daily hospitalizations that result from different durations of strict NPIs under each vaccination scenario. The horizontal dashed lines indicate hypothetical target values of the cumulative number of hospitalizations and the peak daily hospitalizations, where the coloured stars together with the vertical dashed lines indicate the minimum duration of strict NPIs required for each booster scenario to achieve these targets. (C) The trade-off between the cumulative hospitalizations and the peak daily hospitalizations given different durations of strict NPIs.

Figure 7C shows the interrelatedness of the hospitalization targets we selected (dashed lines) for the different vaccination scenarios. The number of days to achieve both targets is similar for the early (blue), faster (orange), and actual NL (black) booster dose scenarios, but for the Canadian average booster dose administration scenario (green) the target total number of hospitalizations is met with many fewer days of strict NPIs (below the horizontal dashed line) than it would take to meet the target peak number of hospitalizations (to the left of the vertical dashed line; Figure 7C).

## 4 Discussion

During the COVID-19 pandemic, Newfoundland and Labrador (NL) had the highest vaccination coverage in Canada (Treble, 2022) and was likely amongst the most vaccinated jurisdictions in the world (see Section 2.1). We used our previously developed and parameterized compartmental model ((Anokye et al, 2026)) to explore vaccination and NPI implementation scenarios. We found that relative to the average vaccination coverage in Canada, the higher vaccination coverage in NL reduced the number of hospitalizations by 519 and the number of deaths by 122 from December 15, 2021 to December 15, 2022 (Figure 6E).

We found that a booster dose rollout that began 35 days earlier (blue bars) or occurred at 150% the rate of the NL booster dose rollout (orange bars), would have reduced the number of hospitalizations by a further 8–14 people and the number of deaths by 3–4 people (Figure 5B). The effectiveness of different vaccination scenarios depends on whether NPIs were implemented. NPIs delay infections, which allows more time to administer vaccines, and can offset the negative impacts of slower vaccination strategies. If no NPIs had been implemented in NL from December 15, 2021 to March 14, 2022 we find that the actual NL booster dose rollout would have performed worse than earlier and faster vaccination scenarios (Figure 5A).

Characteristics of NL were: (i) implementation of some form of NPIs for the first 89 days of Omicron’s spread (Figure 3A; see also (Anokye et al, 2026)); (ii) high primary series coverage when Omicron established (81.3%); and (iii) booster dose administration to around 50% of the population by early February and to 57.3% of the population in the subsequent weeks (Figure 3B). Our results suggest that all of these characteristics were important and acted together to reduce the number of hospitalizations and deaths that occurred in the year following the establishment of the Omicron variant in NL (Figure 6E) for a hypothetical scenario were the BA.1 variant continued to spread (without the emergence of the BA.2 variant) and assuming no waning of immunity.

NPIs are socially and economically disruptive (Clemente-Suárez et al, 2021; Koren and Petö, 2020; Mofijur et al, 2021) and when the Omicron variant established, residents were already fatigued from public health interventions that were implemented since the start of the pandemic two years earlier. A reason why it would have been advantageous to start the booster dose vaccination rollout earlier in NL (blue line) or to administer vaccines at a 1.5 times faster rate (orange line) is that less stringent hospitalization targets, such as no more than 1,700 total hospitalizations or 25 peak daily hospital admissions, might have been met without any implementation of strict NPIs (Figure 7). The actual booster dose administration strategy that occurred in NL was slower and later than our fast and early scenarios, but was still effective, in part, because NPIs were also implemented to delay infections and allow time for more people to receive booster doses before they were infected with the Omicron variant.

A limitation of our work is that we have not stratified infection risk, vaccination coverage, or the probability of hospitalization and death by age, gender, location, ethnicity, socio-economic status, or type of employment. NL’s population has the highest proportion of elderly people in Canada ((Statistics Canada, 2021)), and so we expect that the projected number of hospitalizations and deaths would increase for all scenarios had a realistic age-structure been considered. Immunocompromised people may be particularly vulnerable to infections, even when vaccinated, and are exposed to disproportionate risk when NPIs are suspended, while household members that experience domestic violence are exposed to disproportionate risk when ‘stay-at-home’ requirements are enacted.

An economic analysis might link the cost of NPIs and the cost of slow or late vaccination campaigns to identify an optimal combination of public health measures. Our results contribute to this possible future direction as we use an epidemiologically-based method to quantify the hospitalizations and deaths for different vaccination and NPI scenarios to which the economic considerations might be added. Additionally, we note that investment in vaccination campaigns, particularly ones that occur urgently, can divert resources from other branches of the healthcare system and a holistic analysis would also consider these factors.

Our simulations predicted that NL’s high vaccination coverage prior to the establishment of the Omicron SARS-CoV-2 variant (81.3%) and booster dose vaccination campaign (resulting in 57.3% of NL’s population receiving booster doses) reduced the number of hospitalizations occurring in Newfoundlanders and Labradorians in the next year by 519, and the number of deaths by 122 relative to a counterfactual scenario that implemented vaccination at the average Canadian rate (Figure 6E). These reductions occurred, in part, because NL implemented some form of NPIs during the first 89 days after the Omicron variant established to offset the time that it took to administer booster doses. We find that had the booster dose campaign in NL been implemented at half the actual rate (pink line), NPIs would need to have been enacted for a much longer period to achieve the same hospitalization targets (Figure 7). Our results support the value of vaccinations in protecting populations from severe outcomes of infectious diseases.

## Data Availability

The code and data used for our analysis are publicly available at https://github.com/francisanokye/scenario-modelling-of-vaccination-and-NPIs/

