## Supplementary Information for "High vaccine coverage and non-pharmaceutical interventions decreased hospitalizations and deaths due to the Omicron variant in Newfoundland and Labrador, Canada"

### Supplementary Material

#### S1 Mathematical model

##### S1.1 Model equations

We used the vaccination-stratified SEAIR model described in the main text and previously developed in (Anokye et al, 2026). Here, we provide the complete mathematical formulation and parameter values used in the present analysis. Individuals transition sequentially from the no primary series cohort to the primary series cohort and subsequently to the booster dose cohort; therefore, individuals must complete the primary series before they can receive a booster dose.

The force of infection is

$$\lambda(t) = \beta(t)\frac{\zeta(A_1 + A_2 + A_3) + (I_1 + I_2 + I_3)}{N}, \tag{1}$$

where $\beta(t)$ is the time-varying transmission rate, $\zeta$ is the relative infectiousness of asymptomatic infections, and $N$ is the total population.

Vaccination was incorporated using observed vaccination data (Public Health Agency of Canada, 2024). The time-varying quantities $\nu_2(t)$ and $\nu_3(t)$ specify the scheduled daily numbers of primary-series completions and booster doses, respectively. To preserve non-negativity when the number of eligible individuals becomes small, the realized vaccination flows are

$$f_2(S_1) = \frac{\nu_2(t)S_1}{\nu_2(t) + S_1}, \tag{2}$$

and

$$f_3(V_2) = \frac{\nu_3(t)V_2}{\nu_3(t) + V_2}. \tag{3}$$

Thus, entry into the booster dose cohort requires prior completion of the primary series and is constrained by the number of eligible susceptible individuals in $V_2$.

The system of ordinary differential equations is

$$\textbf{no primary series} \quad \begin{cases} \dfrac{dS_1}{dt} = -\kappa_1\lambda(t)S_1 - f_2(S_1), \\ \dfrac{dE_1}{dt} = \kappa_1\lambda(t)S_1 - \sigma E_1, \\ \dfrac{dI_1}{dt} = \mu\sigma E_1 - \gamma_i I_1, \\ \dfrac{dA_1}{dt} = (1-\mu)\sigma E_1 - \gamma_a A_1, \\ \dfrac{dR_1}{dt} = \gamma_i I_1 + \gamma_a A_1. \end{cases}$$

$$\textbf{primary series} \quad \begin{cases} \dfrac{dV_2}{dt} = f_2(S_1) - \kappa_2\lambda(t)V_2 - f_3(V_2), \\ \dfrac{dE_2}{dt} = \kappa_2\lambda(t)V_2 - \sigma E_2, \\ \dfrac{dI_2}{dt} = \mu\sigma E_2 - \gamma_i I_2, \\ \dfrac{dA_2}{dt} = (1-\mu)\sigma E_2 - \gamma_a A_2, \\ \dfrac{dR_2}{dt} = \gamma_i I_2 + \gamma_a A_2. \end{cases}$$

$$\textbf{booster doses} \begin{cases} \frac{dV_3}{dt} = f_3(V_2) - \kappa_3\lambda(t)V_3, \\ \frac{dE_3}{dt} = \kappa_3\lambda(t)V_3 - \sigma E_3, \\ \frac{dI_3}{dt} = \mu\sigma E_3 - \gamma_i I_3, \\ \frac{dA_3}{dt} = (1-\mu)\sigma E_3 - \gamma_a A_3, \\ \frac{dR_3}{dt} = \gamma_i I_3 + \gamma_a A_3. \end{cases}$$

##### S1.2 Model parameters

Model parameters and their values are summarized in Table S1. The calibration procedure and prior specification are described in (Anokye et al, 2026); only the parameter values used in the present analysis are reported here.

**Table S1**: **Model parameters.** Fixed parameter values were obtained from published estimates or surveillance data. Parameters estimated during model calibration are reported with their 95% credible intervals; the corresponding assumed 95% prior intervals are provided in the Details column.

| Symbol | Definition | Value | Details |
|---|---|---|---|
| **Fixed parameters** | | | |
| $N$ | Total population | 510,550 | (Statistics Canada, 2022) |
| $\mu$ | Proportion symptomatic | 0.676 | (Shang et al, 2022) |
| $\zeta$ | Relative infectiousness of asymptomatic infections ($0 \leq \zeta \leq 1$) | 0.75 | (Johansson et al, 2021) |
| $\kappa_1$ | Relative susceptibility, no primary series | 1.0 | Reference value |
| $\nu_2(t), \nu_3(t)$ | Scheduled daily primary-series completions and booster doses | Vaccination data | (Public Health Agency of Canada, 2024) |
| **Fitted parameters** | | | |
| $\sigma$ | Rate from exposed to infectious | 0.356 days$^{-1}$ (CrI: [0.243, 0.469]) | [1/4, 1/2] days$^{-1}$; (Jansen, 2021) |
| $\gamma_a$ | Recovery rate, asymptomatic | 0.106 days$^{-1}$ (CrI: [0.090, 0.123]) | [1/11, 1/8] days$^{-1}$; (Li et al, 2023) |
| $\gamma_i$ | Recovery rate, symptomatic | 0.144 days$^{-1}$ (CrI: [0.124, 0.164]) | [1/8, 1/6] days$^{-1}$; (Li et al, 2023) |
| $\kappa_2$ | Relative susceptibility, primary series ($0 \leq \kappa_2 \leq 1$) | 0.911 (CrI: [0.862, 0.960]) | [0.85, 0.96]; (Andrews et al, 2022) |
| $\kappa_3$ | Relative susceptibility, booster doses ($0 \leq \kappa_3 \leq 1$) | 0.303 (CrI: [0.229, 0.376]) | [0.20, 0.40]; (Andrews et al, 2022) |
| $\beta(t)$ | Time-varying transmission rate | Time varying | Estimated from seroincidence; see Fig. S1 |

##### S1.3 Transmission rate and control reproduction number

The time-varying transmission rate, $\beta(t)$, was estimated using the calibration framework described in (Anokye et al, 2026). For the present analysis, estimates of $\beta(t)$ for December 15, 2021–March 14, 2022 were used to define the NPI scenarios described in the main text. The corresponding model fit to the seroincidence estimates is shown in Fig. S1.

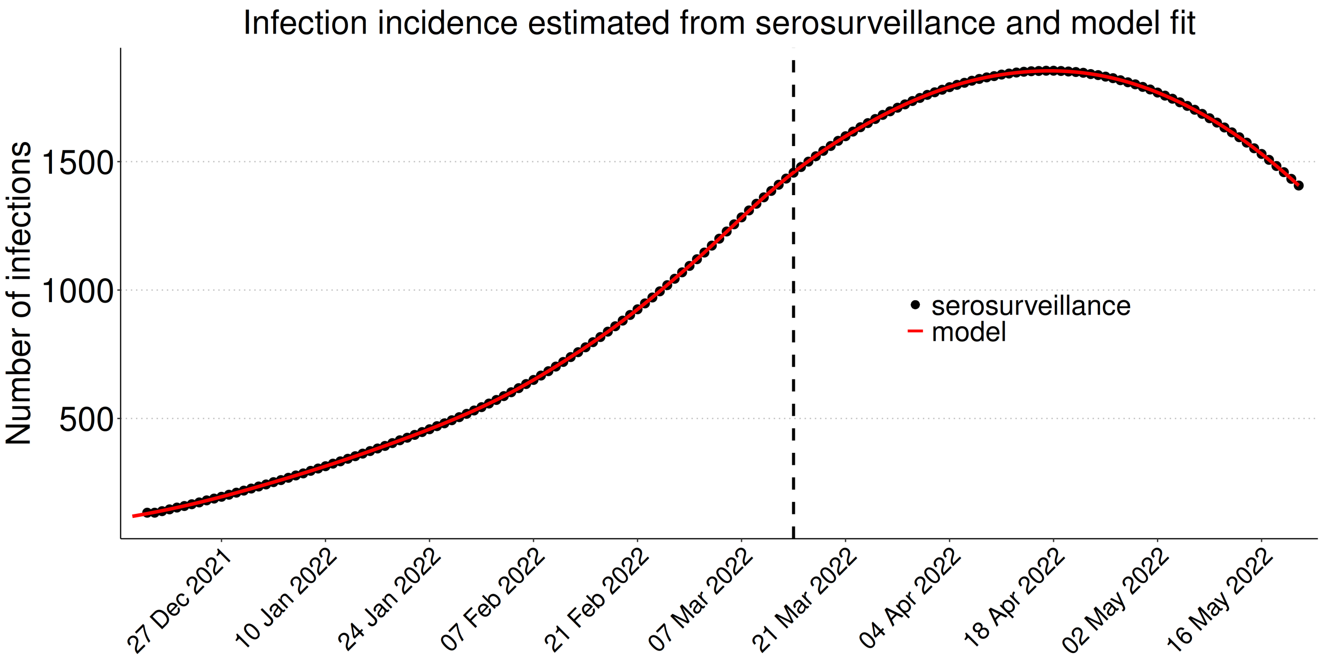


**Fig. S1**: **Model fit to infection incidence estimated from serological prevalence data in Newfoundland and Labrador.** Model-estimated infection incidence (red line; shaded band indicates the 95% confidence interval) is shown against seroincidence estimates (black points). The dashed vertical line indicates March 14, 2022, when the provincial public health emergency was cancelled and the associated Special Measures Orders were repealed.

Following the next-generation matrix approach (Van den Driessche and Watmough, 2002), the control reproduction number was calculated as

$$\mathcal{R}_c(t) = \beta(t) \left( \sum_{j=1}^{3} p_j(t)\kappa_j \right) \left[ \frac{\mu}{\gamma_i} + \frac{(1-\mu)\zeta}{\gamma_a} \right], \tag{4}$$

where

$$p_1(t) = \frac{S_1(t)}{N}, \qquad p_2(t) = \frac{V_2(t)}{N}, \qquad p_3(t) = \frac{V_3(t)}{N}.$$

For comparisons over an interval $[t_a, t_b]$, the mean transmission rate (see Figure S2) was calculated as

$$\bar{\beta}_{[t_a,t_b]} = \frac{1}{t_b - t_a} \int_{t_a}^{t_b} \beta(t)\, dt, \tag{5}$$

and the corresponding reproduction number was obtained by substituting $\bar{\beta}_{[t_a,t_b]}$ for $\beta(t)$ in Eq. 4.

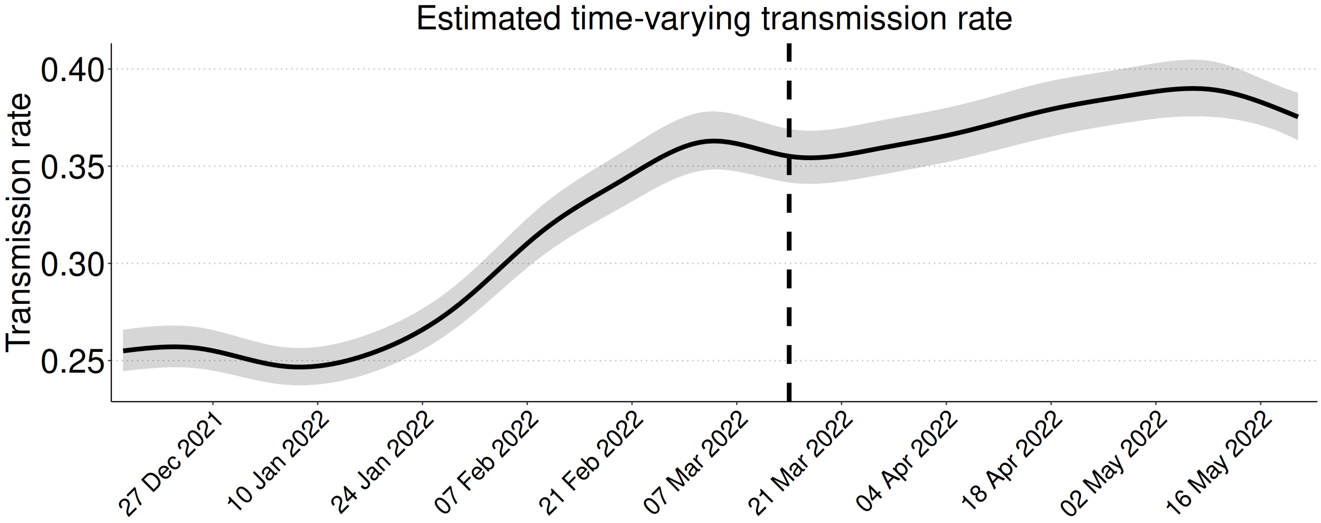


**Fig. S2**: **Estimated time-varying transmission rate of SARS-CoV-2 in Newfoundland and Labrador.** The solid line shows the estimated transmission rate, $\beta(t)$, and the shaded band indicates the 95% confidence interval. The dashed vertical line indicates March 14, 2022, when the provincial public health emergency was cancelled and the associated Special Measures Orders were repealed.

### S2 Additional simulation results

Differences in vaccination speed, timing, and NPI implementation changed the number of infections occurring before booster vaccination (Fig. S3). Faster booster dose administration resulted in fewer infections without booster doses than the actual NL vaccination rollout, whereas slower administration resulted in more infections (Fig. S3A–C). Earlier booster dose administration also reduced infections without booster doses compared with the actual NL vaccination rollout (Fig. S3D–F). Similarly, the actual NL vaccination rollout resulted in fewer infections without booster doses than the Canadian vaccination rate (Fig. S3G–I). Strict NPIs further reduced the cumulative number of infections occurring before booster vaccination across these comparisons.

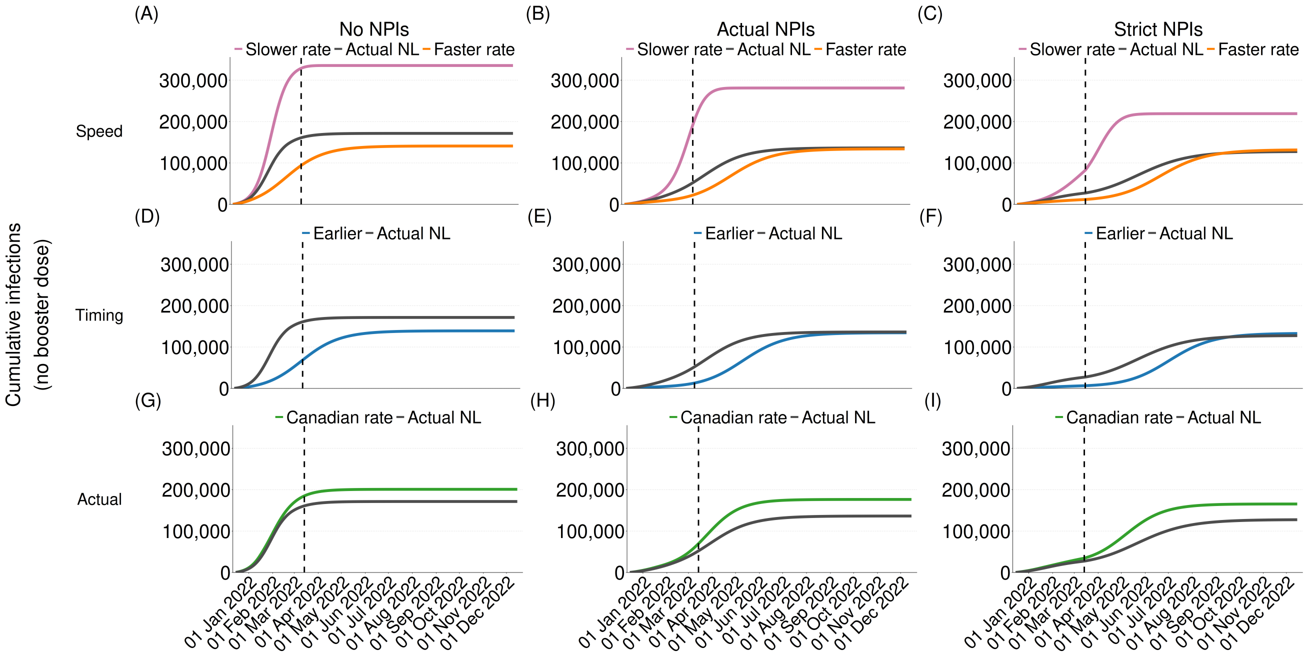


**Fig. S3**: **Cumulative infections without booster doses under different vaccination and NPI scenarios (December 15, 2021–December 15, 2022).** Cumulative infections without booster doses are shown for differences in vaccination speed (A–C), timing (D–F), and the actual NL and Canadian vaccination rollouts (G–I), under no NPIs (left column), actual NPIs (middle column), and strict NPIs (right column). The vertical dashed line indicates the timing of removal of all public health restrictions in NL.

To further examine the differences between the actual NL and Canadian vaccination rollouts, we considered the Canadian vaccination rate with NL initial coverage and the actual NL vaccination rollout with Canadian initial coverage. Differences in both initial vaccination coverage and subsequent vaccination rollout changed the timing of infections occurring before and after booster vaccination (Fig. S4). Strict NPIs delayed infections across all four vaccination scenarios compared with no NPIs, resulting in fewer infections occurring before booster vaccination.

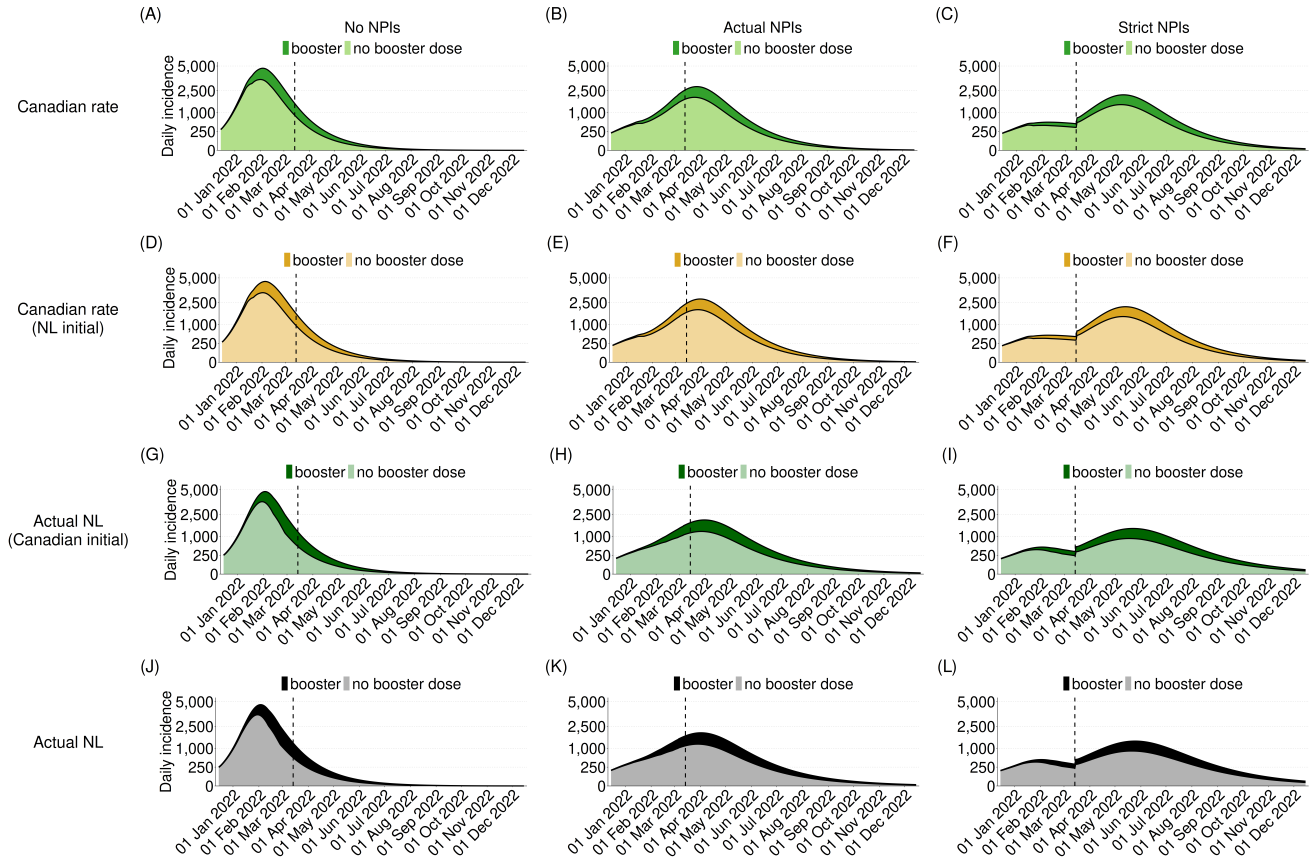


**Fig. S4**: **Temporal infection dynamics under vaccination scenarios combining Canadian and NL initial coverage and vaccination rollouts.** Infections among individuals with no booster dose (light shading) and booster doses (dark shading) are shown under no NPIs (left column), actual NPIs (middle column), and strict NPIs (right column), for the Canadian rate (A–C), Canadian rate with NL initial coverage (D–F), actual NL vaccination rollout with Canadian initial coverage (G–I), and actual NL vaccination rollout (J–L). The vertical dashed line indicates the timing of removal of all public health restrictions in NL. The vertical axis has been square-root transformed to support visualization of the infection trajectories.

The effects of initial vaccination coverage and subsequent vaccination rollout were also reflected in cumulative infections occurring before booster vaccination and in severe outcomes (Fig. S5). Under actual NPIs, the Canadian vaccination rate resulted in 2,004 hospitalizations and 395 deaths. Combining the Canadian vaccination rate with NL initial coverage resulted in 1,801 hospitalizations and 326 deaths, while the actual NL vaccination rollout with Canadian initial coverage resulted in 1,700 hospitalizations and 344 deaths. The actual NL vaccination rollout resulted in 1,485 hospitalizations and 273 deaths (Fig. S5E).

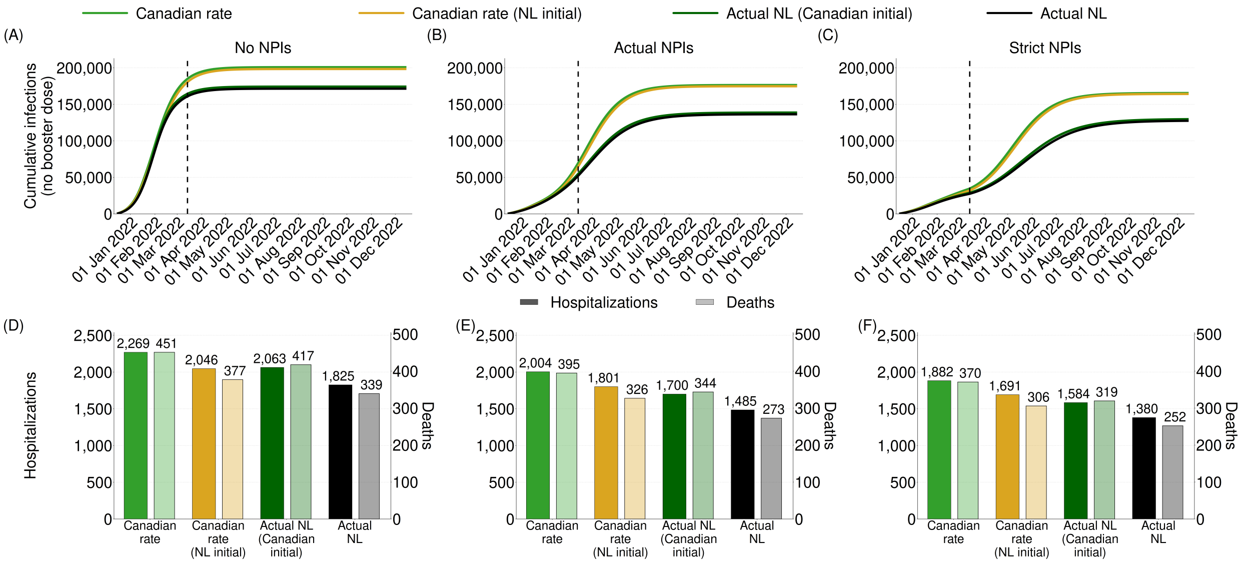


**Fig. S5**: **Cumulative infections and severe outcomes under vaccination scenarios combining Canadian and NL initial coverage and vaccination rollouts.** Cumulative infections among individuals with no booster dose are shown under no NPIs (A), actual NPIs (B), and strict NPIs (C). Total hospitalizations and deaths under the corresponding vaccination scenarios are shown in D–F. The vertical dashed line indicates the timing of removal of all public health restrictions in NL.

The differences among the four vaccination scenarios were most apparent in cumulative infections occurring before booster vaccination (Fig. S6A–C). Cumulative infections after booster vaccination were similar across the four vaccination scenarios under no NPIs and actual NPIs, with somewhat greater differences under strict NPIs (Fig. S6D–F). Across the NPI scenarios, vaccination scenarios using the actual NL vaccination rollout resulted in fewer infections before booster vaccination than the corresponding scenarios using the Canadian vaccination rate.

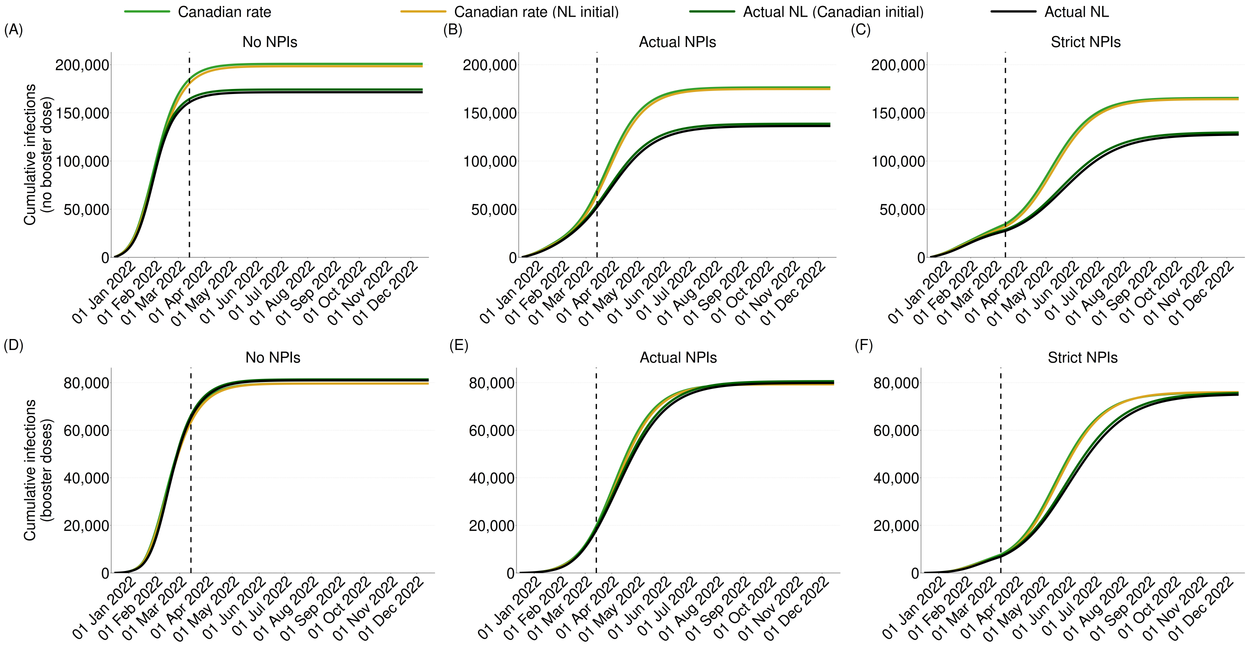


**Fig. S6**: **Cumulative infections by booster dose status under the four vaccination scenarios.** Cumulative infections among individuals with no booster dose (A–C) and booster doses (D–F) are shown under no NPIs (left column), actual NPIs (middle column), and strict NPIs (right column), for the Canadian rate, Canadian rate with NL initial coverage, actual NL vaccination rollout with Canadian initial coverage, and actual NL vaccination rollout. The vertical dashed line indicates the timing of removal of all public health restrictions in NL.

The vaccination trajectories show how initial vaccination coverage and subsequent booster dose administration differed among the four scenarios (Fig. S7A–C). Scenarios with NL initial coverage began with fewer individuals without a primary series than the corresponding scenarios with Canadian initial coverage (Fig. S7A), while scenarios using the actual NL vaccination rollout reached greater final booster dose coverage than those using the Canadian vaccination rate (Fig. S7C). These differences corresponded to differences in hospitalizations and deaths across the four vaccination scenarios (Fig. S7D–F).

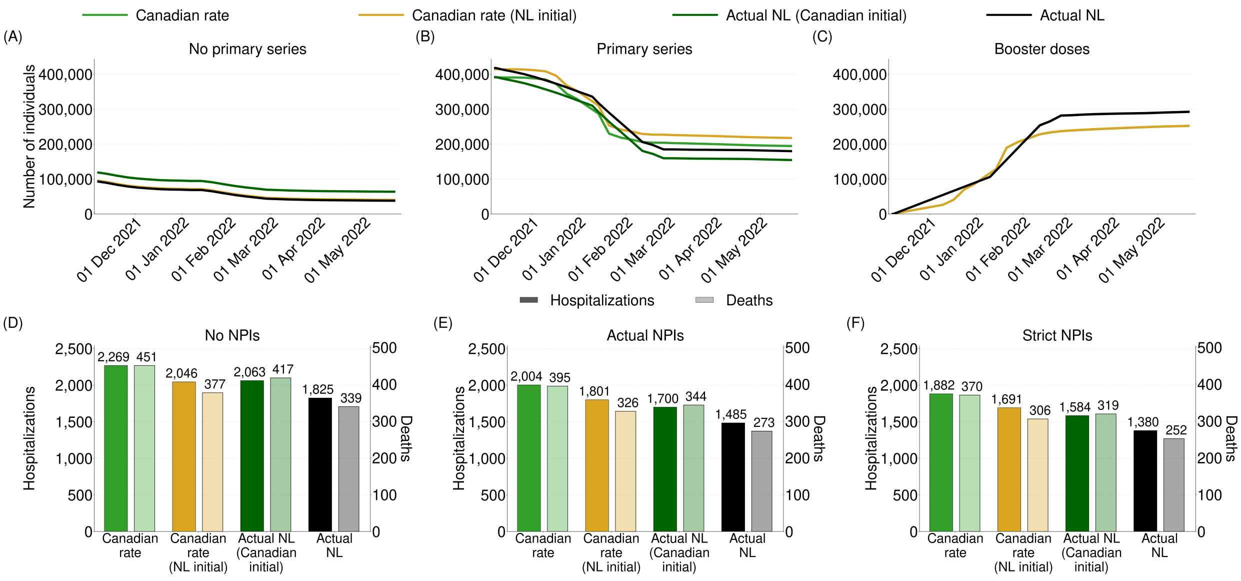


**Fig. S7**: **Vaccination coverage and severe outcomes under vaccination scenarios combining Canadian and NL initial coverage and vaccination rollouts.** The numbers of individuals with no primary series (A), primary series (B), and booster doses (C) are shown for the Canadian rate, Canadian rate with NL initial coverage, actual NL vaccination rollout with Canadian initial coverage, and actual NL vaccination rollout. Total hospitalizations and deaths are shown under no NPIs (D), actual NPIs (E), and strict NPIs (F).
